# Variability of Tissue Mechanical Response in *Sus Domesticus* Porcine Models from *in vivo* to *ex vivo* Conditions

**DOI:** 10.1101/2022.05.05.22274734

**Authors:** Faizan A. Malik, Bradley A. Drahos, Amer Safdari, Mark V. Mazzeo, Jack E. Norfleet, Robert M. Sweet, Timothy M. Kowalewski

## Abstract

Healthcare simulators have been demonstrated to be a valuable resource for training several technical and nontechnical skills. A gap in the fidelity of tissues has been acknowledged as a barrier to application for current simulators; especially for interventional procedures. Inaccurate or unrealistic mechanical response of a simulated tissue to a given surgical tool motion may result in negative training transfer and/or prevents the “suspension of disbelief” necessary for a trainee to engage in the activity. Thus, where it is relevant to training outcomes, there should be an effort to create healthcare simulators with simulated tissue mechanical responses that match or represent those of biological tissues. Historically, this data is most often gathered from preserved (post mortem) tissue; however, there is a concern that the mechanical properties of preserved tissue, that lacks blood flow, may lack adequate accuracy to provide the necessary training efficacy of simulators. This work explores the effect of the “state” of the tissue testing status on liver and peritoneal tissue by using a customized handheld grasper to measure the mechanical responses of representative porcine (*Sus domesticus*) tissues in N=5 animals across five test conditions: in vivo, postmortem (in-situ), ex vivo (immediately removed from fresh porcine cadaver), post-refrigeration, and post-freeze-thaw cycle spanning up to 72 hours after death. No statistically significant difference was observed in the mechanical responses due to grasping between *in vivo* and post-freeze conditions for porcine liver and peritoneum tissue samples (p = 0.05 for derived stiffness at grasping force values F = 5N and 6.5N). Furthermore, variance between *in vivo* and *post-thaw* conditions *within* each animal, was comparable to the variance of the *in vivo* condition *between* animals. Thus, the use of thawed, preserved tissue for the further study and emulation of mechanical perturbation of the liver and peritoneum can be considered.

## Introduction

The lack of accurate tissue mechanical behavior is a well-recognized barrier to the wide-spread adoption of simulated analogues for moderate to complex interventional healthcare procedures. The state-of-the-art approach for improving the mechanical characteristics of medical simulators is to model them after data derived from existing literature on tissue biomechanics. A potential limitation is that much of the existing tissue characterization data is measured outside of the tissue’s natural environment, often-times long after the host has died. One concern is that by using this pre-existing data based on ex vivo tissue, physical and virtual analogues will simply behave more cadaveric rather than mimicking the mechanical properties and behaviors of perfused tissue. Little is currently known about how contemporary biomechanical characterization techniques truly match the properties of the tissues in vivo. If this discrepancy was further studied and characterized, it could provide evidence either in support of using existing, widespread ex vivo data as suitable for the development of simulated healthcare models or contrary evidence, emphasizing the need for in vivo data.

### Prior Work

One of the earliest and most notable direct comparisons between in vivo and ex vivo tissue was conducted by Rosen et al. in 2008 (1). The purpose of this study was to characterize the changes in tissue mechanical properties measured ex vivo to their in vivo state to aid in the development of more mechanically realistic medical simulators; that is, a simulated tissue’s force-motion profile should resemble that of its targeted living tissue. During this study, Rosen et al. developed a handheld grasping device (MEG) which could apply compressive loads to tissue and monitor applied stresses and strains. This device was designed such that it could gather data in vivo which could then be compared to results from testing tissue ex vivo using a uniaxial compressive fixture (MTS). Both sets of data were fit with a phenomenological modeling approach and the key finding was that properties of the tissue had a noticeable difference from in vivo to ex vivo for most organs. It was found that stresses were higher at a set strain in vivo. While these results were a key finding, this study noted its limitation to phenomenological models and measurements (i.e., curve fitting) and did not reveal fundamental underlying mechanisms that caused the measured changes. However, for medical simulators such phenomenological measurements or models may suffice as a basic means to measure whether a simulated physical tissue matches its real living target to in a quantifiable manner.

A similar study, conducted by Mazza et al., analyzed mechanical properties of the human cervix in vivo and ex vivo both before and after a hysterectomy procedure (2). For this study a stiffness parameter was used to characterize tissue mechanical properties. They were ultimately unable to determine any statistically significant difference in stiffness between in vivo and ex vivo cervical tissue from their aspiration experiment. However, the authors did cite that the lack of statistical significance could potentially be due to large variability in the data, thus further analysis could yield differing results.

Around the same time as the previous studies, Ocal et al. verified a common assumption that the longer tissue is preserved ex vivo, the stiffer and more viscous it becomes through testing of bovine liver tissue (3). The primary result from this study was the determination that as tissue was stored postmortem, the loss and relaxation moduli of the tissue increased. This indicates that as the tissue is stored over time the tissue becomes stiffer and more viscous. It is important to note that a key limitation of this study was a lack of in vivo testing of the tissue or a trial immediately postmortem.

For this study, a primary source of concern was whether tissue damage would affect tissue mechanical response. Kerdok et al. analyzed the effects of perfusion on mechanical properties of porcine liver (4). The primary conclusion from Kerdok’s study is that there was a statistically significant difference between in vivo tissue and ex vivo post perfused tissue. Both sets of tissue had similar viscoelastic properties, with the minimal differences being attributed to inconsistent perfusion pressures and concentrations. Ultimately, the tissue was sent to histologists which reported little to no damage; thus, the perfusion kept the tissue mechanically preserved.

Brouwer et al. in 2001 analyzed mechanical property characterization between in vivo and ex vivo porcine tissue to provide more accurate haptics for surgical simulation (5). This study made use of an indentation device that could be used to compress tissues that were too fragile for tensile tests. The resulting data was fit to exponential curves with high fit and it was determined that a discrepancy existed among fit parameters between in vivo and ex vivo tissue. This discrepancy was attributed to changes in boundary conditions from in vivo to ex vivo. It should be noted that a limitation of this study is the lack of relatability between in vivo and ex vivo experiments.

Samur et al. (6) tested in vivo porcine liver using an indentation instrument guided by laparoscopic ports to minimize invasiveness. The data was used in an inverse finite element simulation to extract material properties, and thus did not directly categorize mechanical response.

Lim et al. (7) introduces a robotic measurement system to obtain the mechanical response of tissue. This study, however, only utilized in-situ cadaveric tissue, and developed various tissue models with the assumptions of incompressible, homogenous, and isotropic tissue. While these assumptions are reasonable for some solid tissues like the liver, these assumptions are not realistic for “hollow” tissues like the bowel or bladder.

Prior work in this project published by Safdari in 2019 (8) investigated characterizing tissue mechanical response in different states. While the study only analyzed the tissues in the *postmortem*, to *ex vivo*, to *post-freeze* tissue states, it found that *post-freeze* liver and spleen tissue exhibited measurable, statistically significant lower stiffnesses compared to *postmortem* tissue. The study introduced techniques to analyze the force-strain data (mechanical response) of porcine tissue under multiple conditions; this work aims to extend and optimize those techniques, as well as incorporate the critical *in vivo* tissue state into the analysis. The study, like this work, used a manual, human-operated grasper device. While this does limit repeatability due to inherent human variances in strain rates and force, it does ensure that the tissues are tested within realistic human viscoelastic regimes.

### Research Objective and Contributions of this Paper

This study aims to measure the change in mechanical responses encountered in typical surgical grasping in porcine models between tissue in its native living (in vivo) state and four deceased states: in vivo, post-mortem, ex vivo, post-refrigeration, and post-freeze. Descriptions for experimental conditions of each state appear in Table 1. The target tissues were limited to intact whole organs or complexes such as liver, spleen, peritoneal tissue (peritoneum), and lung. For each of these tissues, a manual grasping motion following a repeated, audio cued timing cadence was applied to the tissue, and the resulting force-displacement recordings were used to infer and analyze the tissue’s stiffness at set compressive loads of F=5N and 6.5N, that fell in a common range observed during testing. Measured changes in inferred tissue stiffness across all five states (from in vivo to thawed) within a given animal were compared to measurements between all N=5 animals.

**Table 1:** Tissue Testing Conditions.

| Condition | Tissue State | Time After Death [hr.] | in/ex situ |
| --- | --- | --- | --- |
| 1 | in vivo | N/A | in-situ |
| 2 | postmortem | 1-2 | in-situ |
| 3 | ex vivo | 5-6 | ex-situ |
| 4 | post-refrigeration | 24-36 | ex-situ |
| 5 | post-freeze | 48-72 | ex-situ |

Table 2 summarizes and compares the work discussed in the introduction.

**Table 2:** Comparison of Prior Work.

| Reference | *in vivo* | *in-situ* | *ex vivo* | Modeling | Test Specimen | Test Type |
| --- | --- | --- | --- | --- | --- | --- |
| Rosen et al. (1) | x |  | x |  | Porcine organs | Tension, indentation |
| Mazza et al. (2) | x |  | x |  | Human cervix | aspiration |
| Ocal (3) |  |  | x | maxwell-voigt | Porcine liver | Impact, stress relaxation |
| Kerdok et al. (4) | x |  | x | maxwell-voigt | Porcine liver | indentation |
| Brouwer et al. (5) | x |  | x |  | Porcine organs | Tension, indentation |
| Lim et al. (7) |  | x |  | quasi-linear viscoelastic | Human cadaver<br>stomach | Indentation |
| Samur et al. (6) | x |  |  | mooney-rivlin, finite<br>element | Porcine liver | Indentation, stress<br>relaxation |
| Safdari et al. (8) |  | x | x | mooney-rivlin/ogden<br>finite element and<br>model free | Porcine organs | Indentation with hand-<br>held grasper device |
| Current Study | x | x | x |  | Porcine organs | Indentation with hand-<br>held grasper device |

## Materials and Methods

### Approval and Ethics Statement

*This study was carried out in accordance with the recommendations in the Guide for the Care and Use of Laboratory Animals of the National Institutes of Health. The protocol was approved by the Institutional Animal Care and Use Committee of the University of Minnesota (Protocol Number: 1805-35922A); as well as the USAMRMC Animal Care and Use Review Office (ACURO) under Award Number W911NF-14-2-0035. All surgery was performed under anesthesia, and all efforts were made to minimize suffering*.

### Porcine Models and Tissue Handling

An overview of the study population is shown in Table 3 below.

**Table 3:** Study Population Overview.

| Condition | Gender | Session Start<br>Date | Time of<br>Death | Body<br>Temp (°C) | Weight<br>(kg) | Date of Birth |
| --- | --- | --- | --- | --- | --- | --- |
| 1 | F | 2019/15/07 | 12:40 PM | 35.6 | 76.4 | 3/7/19 |
| 2 | F | 2019/24/07 | 12:40 PM | 36.9 | 80.4 | 3/7/19 |
| 3 | F | 2019/07/08 | 9:29 AM | 38.6 | 71.2 | 4/3/19 |
| 4 | F | 2019/18/09 | 11:59 AM | 38.1 | 79.2 | 5/23/19 |
| 5 | F | 2019/23/10 | 11:40 AM | 38.7 | 83.0 | 5/23/19 |

Only Yorkshire-X type pigs were used

### Anesthesia

The animal was given a dose of 5-7 mg/kg Telazol i.m., an ear vein catheter was started and the swine was given an additional 5-7 mg/kg methohexital i.v. and given 0.9% saline through the ear vein during the procedure. The animal was intubated and maintained on isoflurane >1.2 MAC for the remainder of the procedure. An arterial line was placed in a branch of the femoral artery to monitor blood pressure.

A medial sternotomy was performed using electrocautery and a sternal saw. The sternum was retracted and the pericardium was removed or sutured in the corners to produce a pericardial cradle. Lipovenous or other combinations of omega-3 fatty acids were administered to promote viability in-vitro in some subjects.

### Euthanasia

After data collection was complete, the heart was isolated for separate, unrelated in vitro research in order to maximize the use of the animal. The swine remained under a deep plane of anesthesia during all procedures prior to euthanasia.

∼30,000 units of heparin were administered via i.v.; an aortic root cannula was sutured into the ascending aorta; the inferior vena cava and ascending aorta were clamped and high potassium cardioplegia was administered via the aortic root cannula under 150 mm/hg pressure. The superior vena cava was clamped and a small incision was made in the pulmonary artery. The heart was arrested before being excised for other unrelated studies.

### Facilities

The animal experiments were conducted at a dedicated operating facility (Visible Heart Lab) where each animal was used for additional unrelated scientific studies regarding the heart. Immediately after the postmortem studies, the tissues were taken to a separate nearby facility (Wet Lab). The procedure is shown in Fig 1.

**Fig 1.**
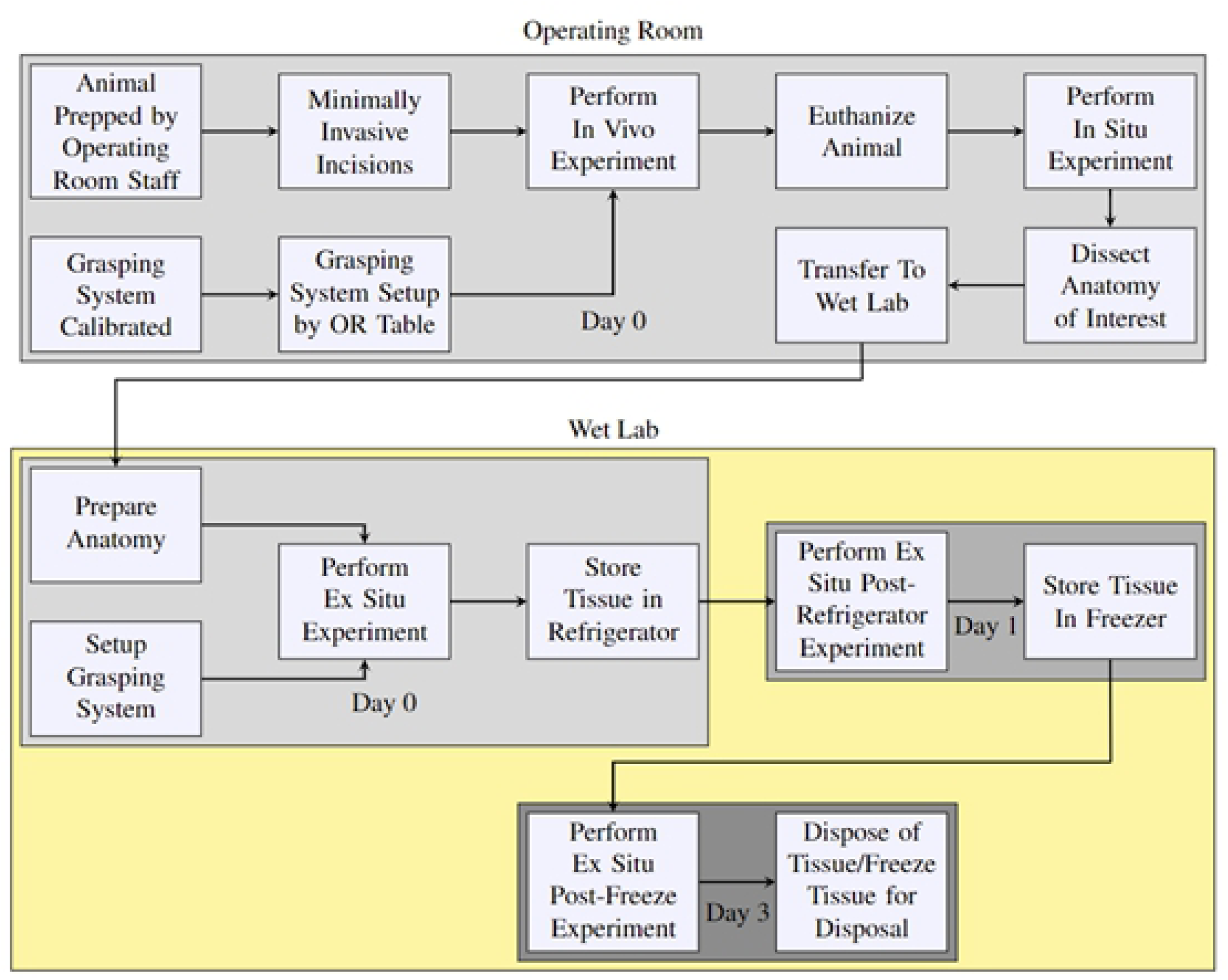
Tissue handling and testing overview.

### In Vivo and Postmortem

An incision was made starting in the abdominal cavity (to access the liver, spleen, and “peritoneal” tissues), and then later expanded to the chest cavity (to access the lung and aorta). The body temperature of the animal was measured via a rectal thermometer and recorded.

After the data collection process on the desired tissues was complete, the animal was euthanized and its heart was extracted using the procedure above. Immediately after euthanasia (time was noted), data collection continued on the same tissues that were tested during *in vivo* conditions.

### Ex Vivo, Refrigeration, and Freezing

After postmortem testing, the tissues were extracted from the carcass and placed in a plastic bag with 25 mL of 1x PBS/pen-strep antibiotic solution. Due to its larger size, the liver was placed in a bag with 75 mL of the same solution.

The plastic bags containing the tissues were held in water baths at 37.5 degrees Celsius until data was collected for the *ex vivo* stage. After *ex vivo* data collection, the tissues were replaced in their respective plastic bags (still containing the antibiotic solution), and refrigerated at approximately 4 degrees Celsius.

After approximately 24 hours of refrigeration, the bags containing the tissues were removed from the refrigerator and reheated in a water bath at 37.5 to 38 degrees C for 2-4 hours. The tissues remained in the antibiotic solution until they were temporarily removed for data collection. After data collection was completed, the tissues were replaced in plastic bags. As the bags became saturated with fluids from the tissue, the bags were replaced as was the antibiotic solution.

The tissues were then placed in a freezer at -18 degrees C for approximately 48 hours, after which they were reheated in a water bath at 38 degrees C for 4-5 hours. Data was collected from the tissues for a final time, after which the tissues were disposed.

The tissue testing conditions are summarized in Table 1.

Actual temperatures were measured at all stages of the protocol stated above; measured temperatures for the liver and peritoneum are shown in S1 Fig.

### Grasping Device Design

The device used for all data collection was a scissor-like tissue grasping device designed by the Medical Robotics and Devices lab. A detailed discussion of the grasping device and its measurement performance is described elsewhere (8). The device contains two opposing load cells (TAL220B, HT Sensor Technology) in a scissor-like fashion, with 3D-printed hemispherical plastic tips serving as the grasping surface and a quadrature rotary encoder (AMT102-V, CUI Inc.) to measure the angle of the arms as the device opens to monitor tissue thickness and overall displacement. A depiction of the grasper device is shown in Fig 2.

**Fig 2.**
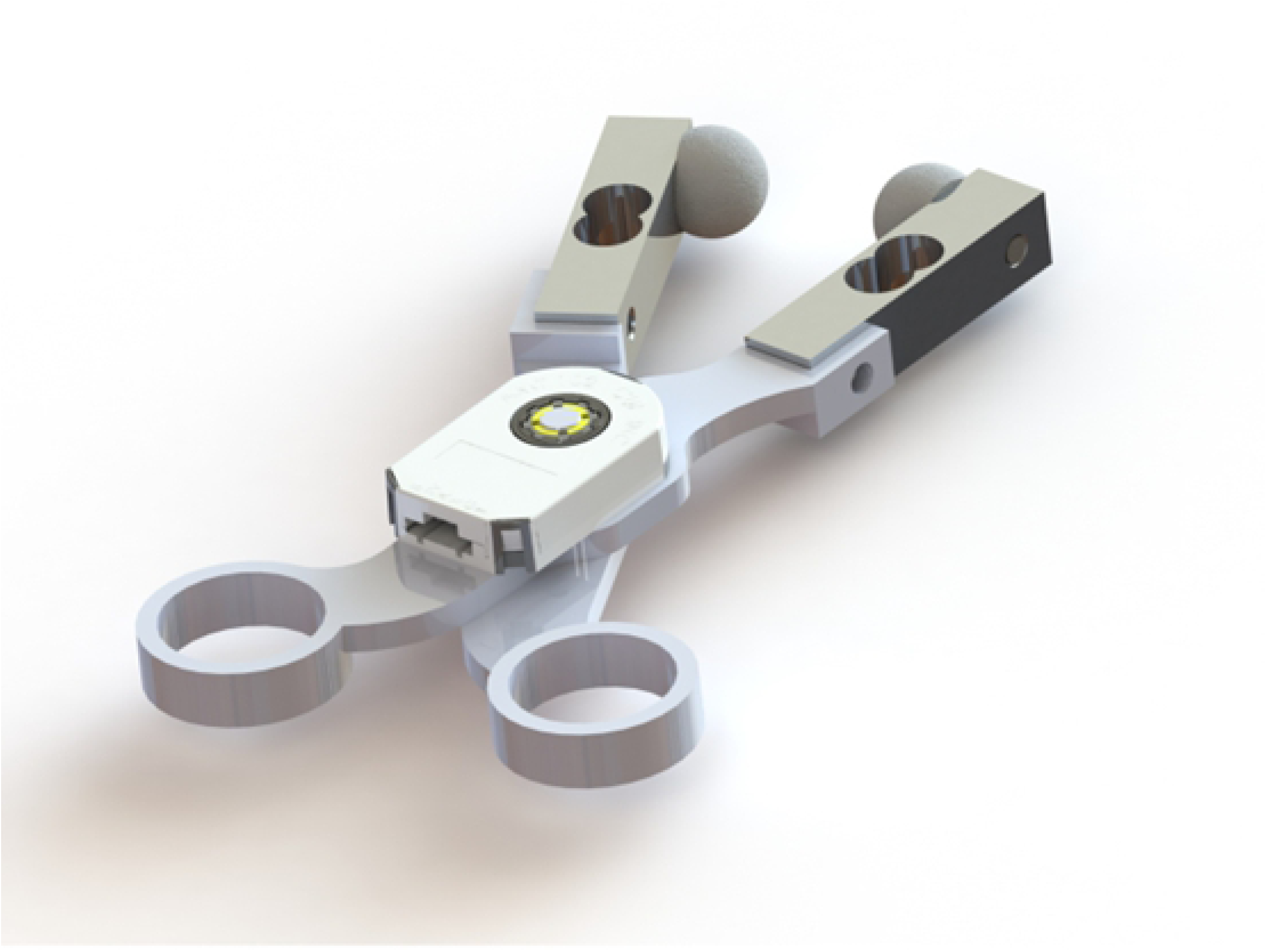
Rendered image of the handheld grasper device.

The computer running the Scissors Console application utilized NI DAQMX drivers to communicate with the DAQ and acquire the voltage and encoder data at a frequency of 1 kHz. The Scissors Console application automatically stored the data in CSV text files of the user’s choosing, and also displayed a live graph of the data. Fig 3 shows how the data is collected from the sensors.

**Fig 3.**
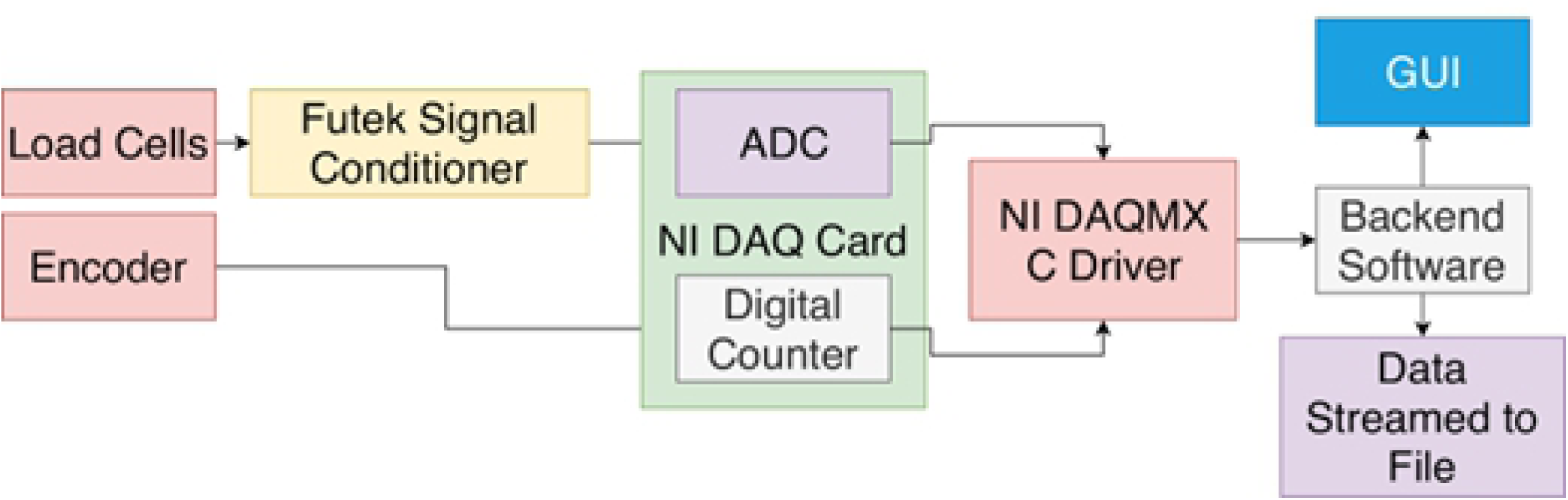
The flow of data from the grasper sensors.

### Grasping Device Operation

#### Setup

The grasping device was placed into a medical glove such that the load cells are placed into the fingers of the glove: the top jaw went into the glove’s “index finger” and the lower jaw was placed in the glove’s “pinkie finger” (most ulnar, smallest finger). The remaining fingers of the glove were taped against the device in order to prevent interference during testing, as seen in Fig 4.

**Fig 4.**
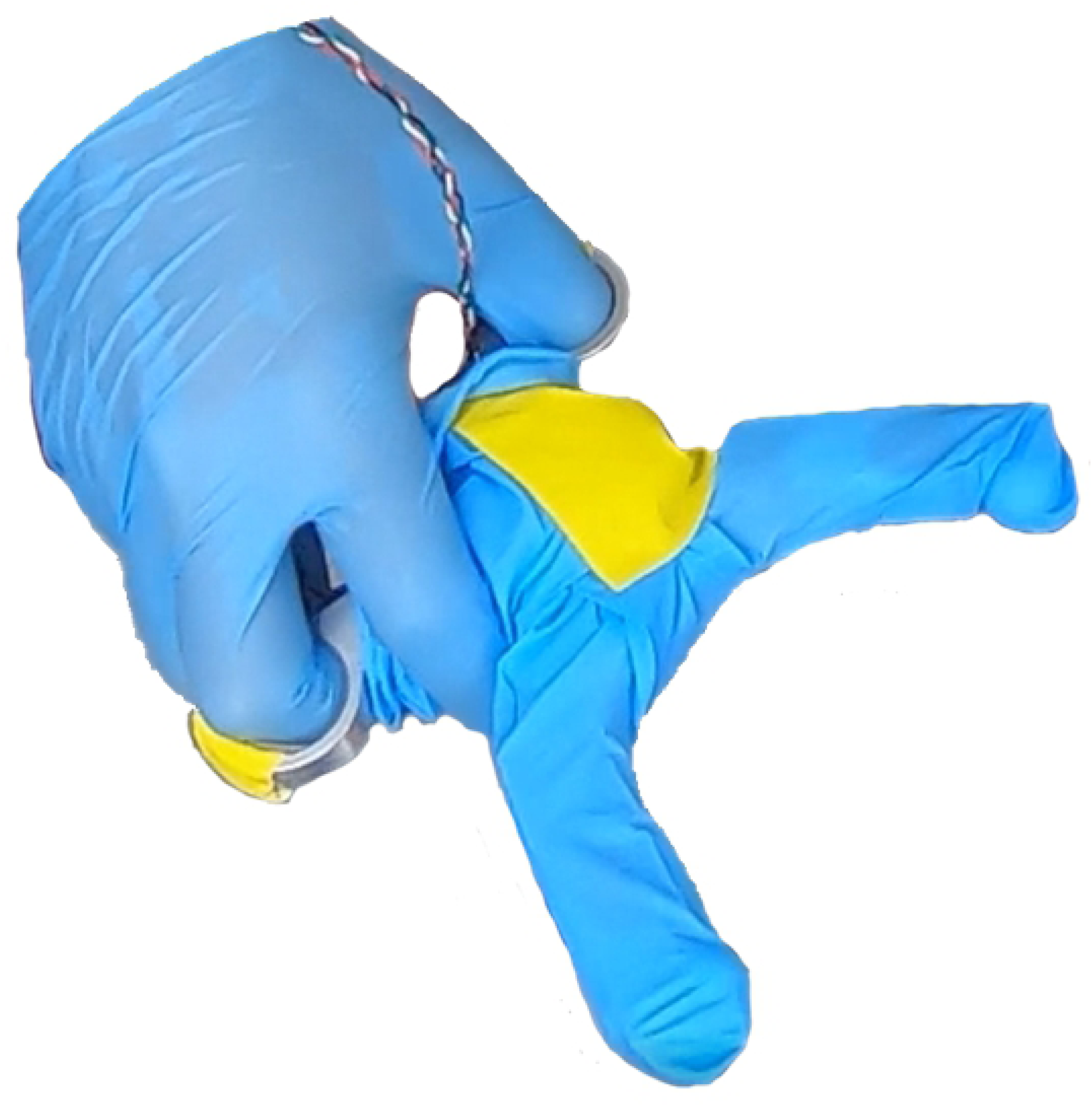
A picture of the grasper device in gloves, being held by a gloved hand.

The grasping device was connected to a National Instruments DAQ, which was then connected to a computer. The computer ran the “Scissors Console” application, which sampled the voltage and encoder data at 1000 Hz, and stored the data in CSV files defined by the user. The application also displayed a real-time graph of the data, as shown in Drahos et al.(9). Additionally, the application produced a series of audio tones at fixed time intervals to foster consistent grasp timings; a “grasp tone” was played for 1.5 seconds, with a delay of 1.7 seconds until the next tone.

## Results

Fig 5 shows changes in stiffness measured in the Liver for the postmortem, ex vivo, and post-refrigeration states compared to the in vivo state were statistically significant. Stiffness also measurably increased under the post-thaw state; however, the change was not statistically significant. The remaining discussion compares the in vivo and post-thaw states.

**Fig 5.**
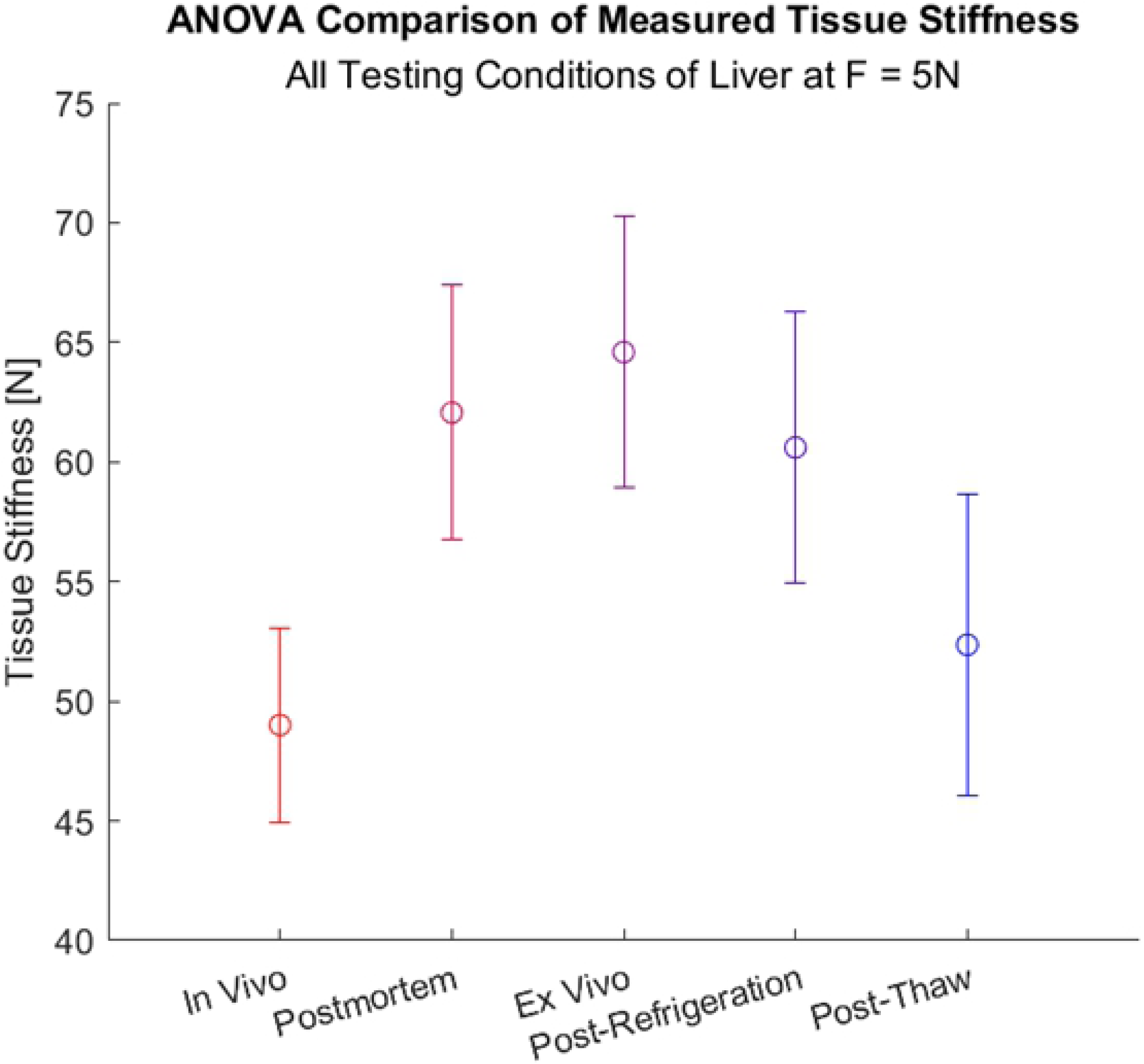
ANOVA comparison of measured stiffness of the liver at F=5N. This plot shows the ANOVA confidence intervals (at p = 0.05) for the Liver, comparing the in vivo tissue state to the other tissue states (postmortem, ex vivo, post-refrigeration, and post-thaw).

Fig 6(A) and (B) show that in vivo liver stiffnesses were fairly consistent across patients; however, post-thaw stiffnesses varied substantially (generally increased stiffness). Peritoneal stiffnesses shown in Fig 6(C) and (D) varied substantially from patient to patient; however, the post-thaw stiffness for each patient was generally consistent with the in vivo data. The average change in stiffness from in vivo to post-thaw conditions for each tissue were relatively small compared to the measured tissue stiffnesses, as shown by the “average delta”.

**Fig 6.**
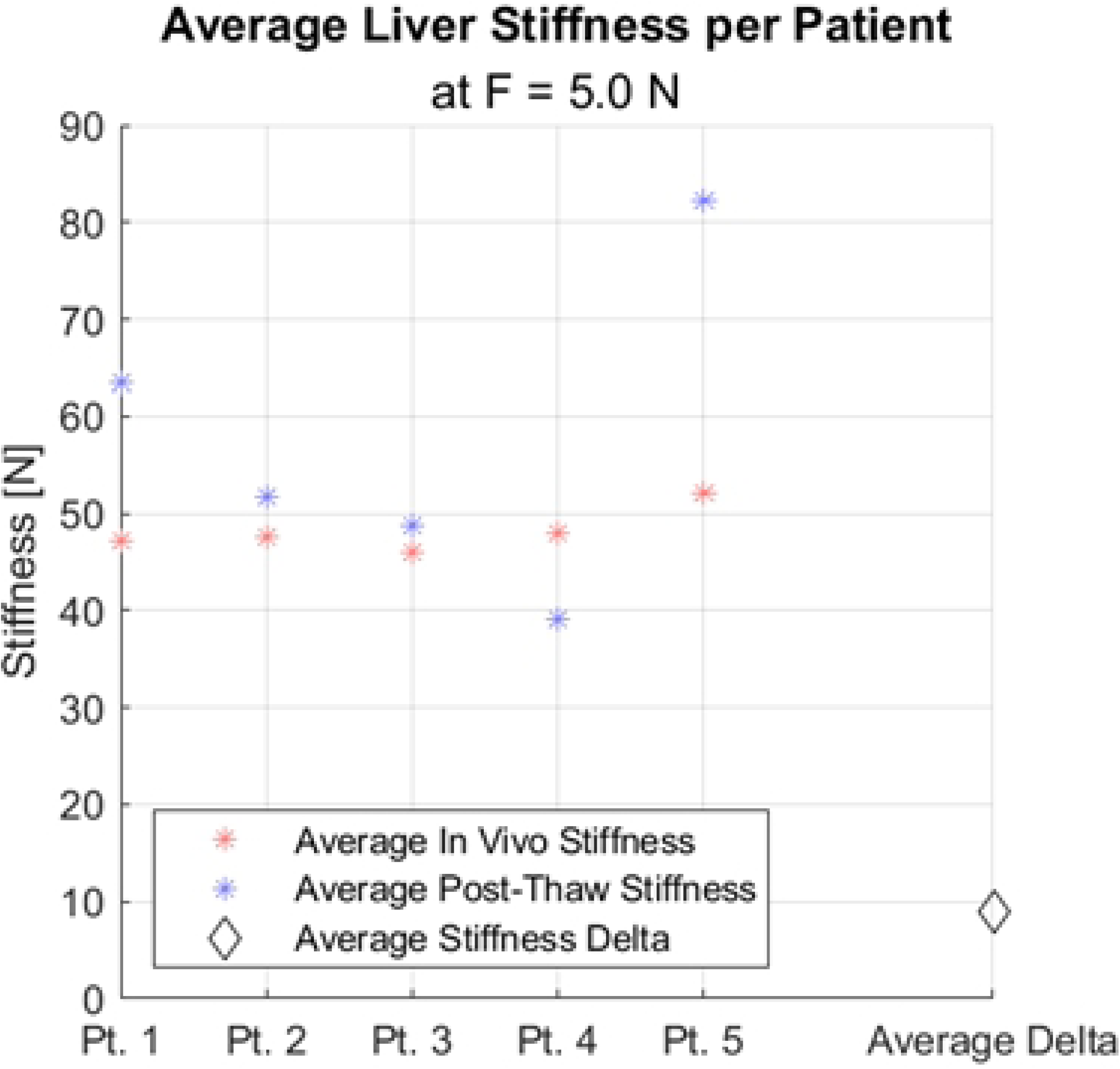
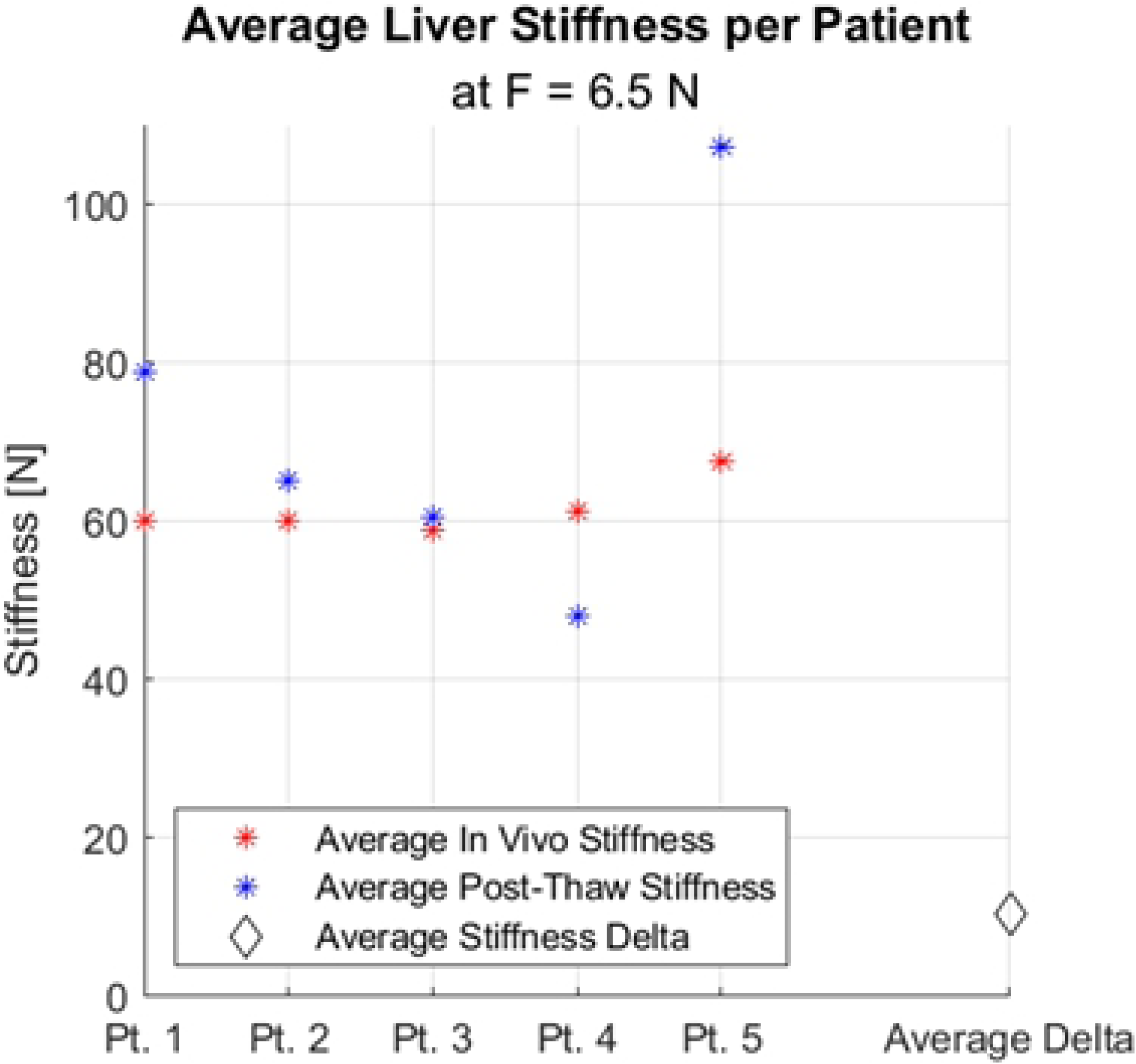
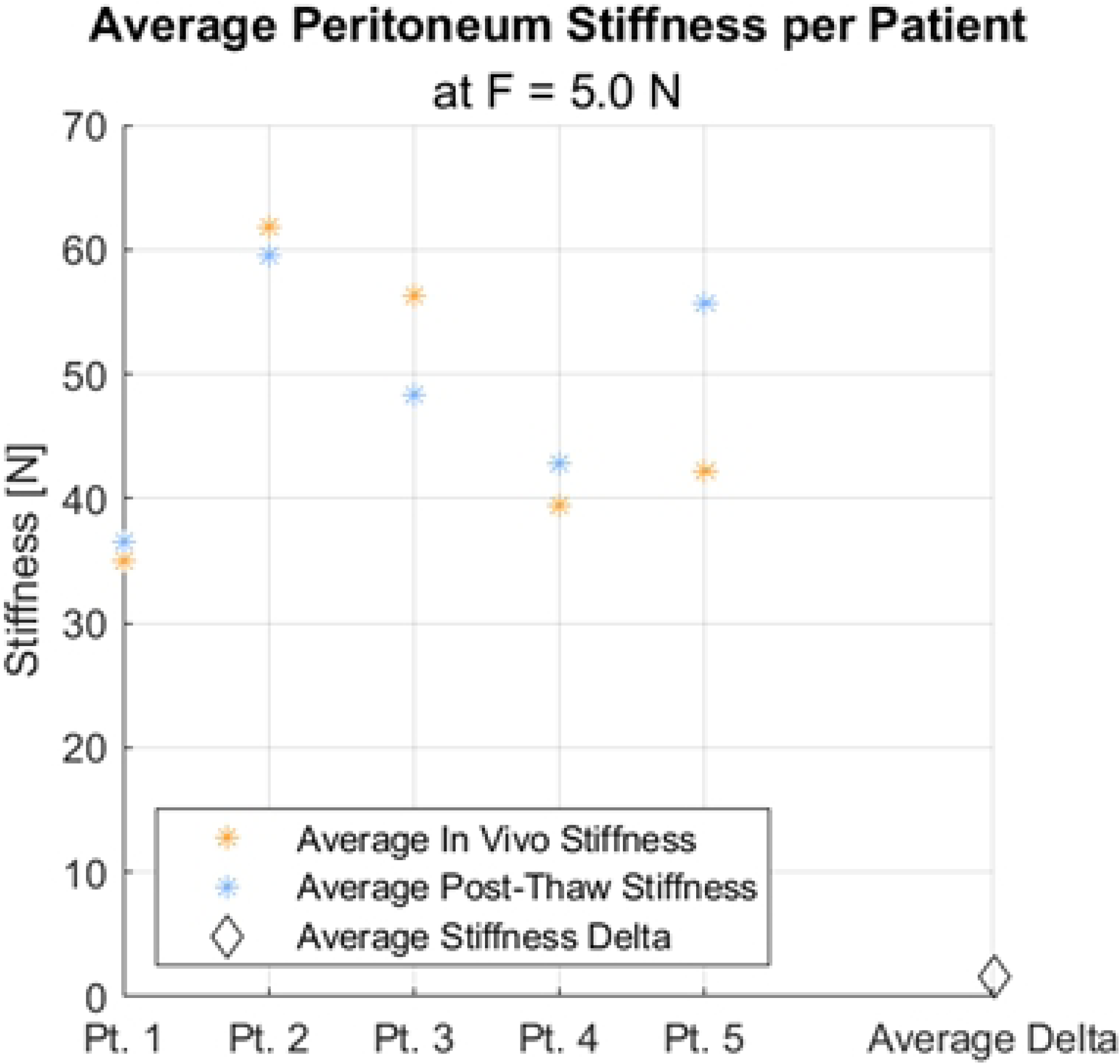
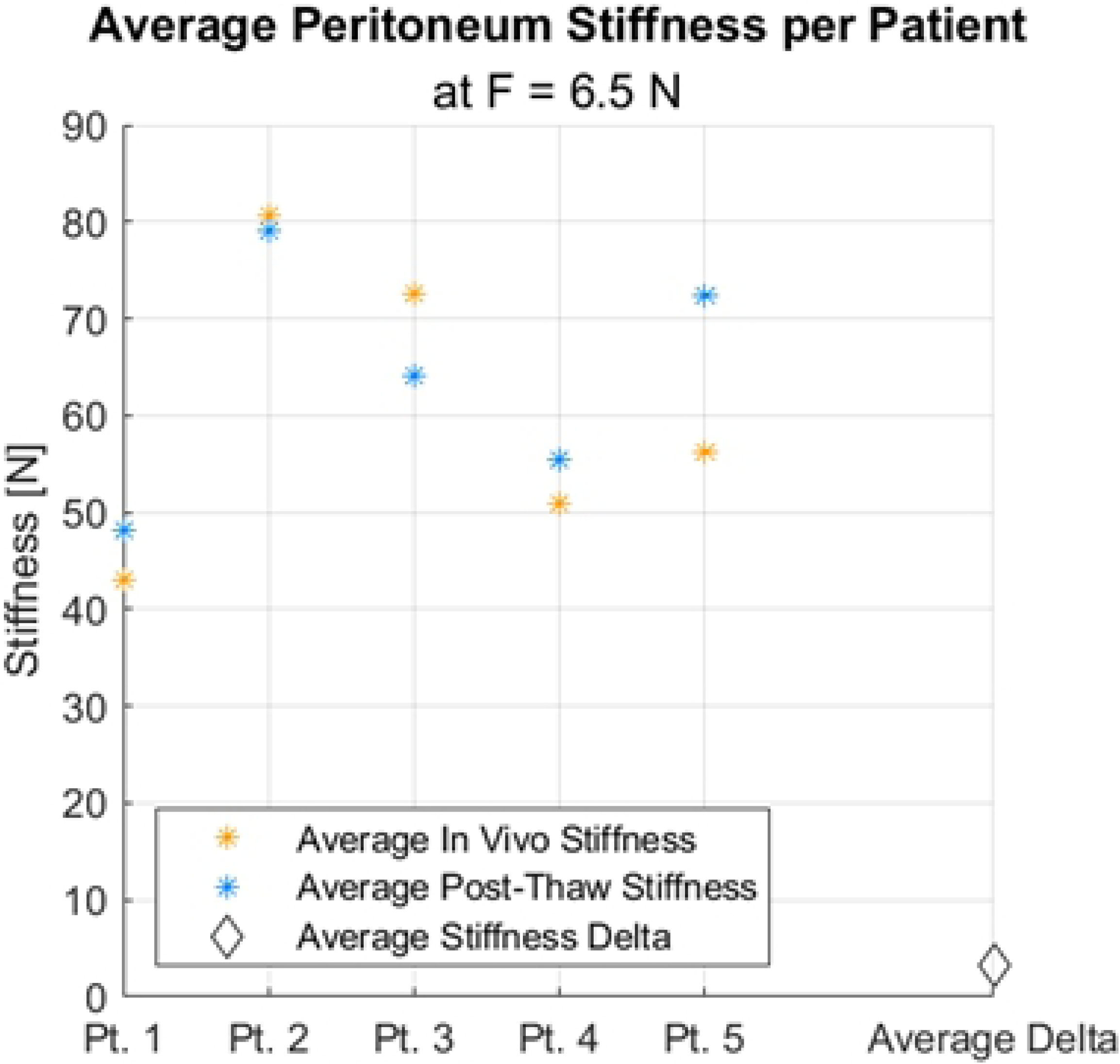
Average measured tissue stiffnesses during in vivo and post thaw, per patient. (A) Liver at F=5N. (B) Liver at F=6.5N. (C) Peritoneum at F=5N. (D) Peritoneum at F=6.5N.

Fig 7 shows that the changes in stiffness from in vivo to post-thaw conditions were not statistically significant. The variability in stiffness of the tissue under a particular condition (in vivo or post-thaw) was comparable with or exceeded the average change in stiffness from changing the tissue condition from in vivo to post-thaw.

**Fig 7.**
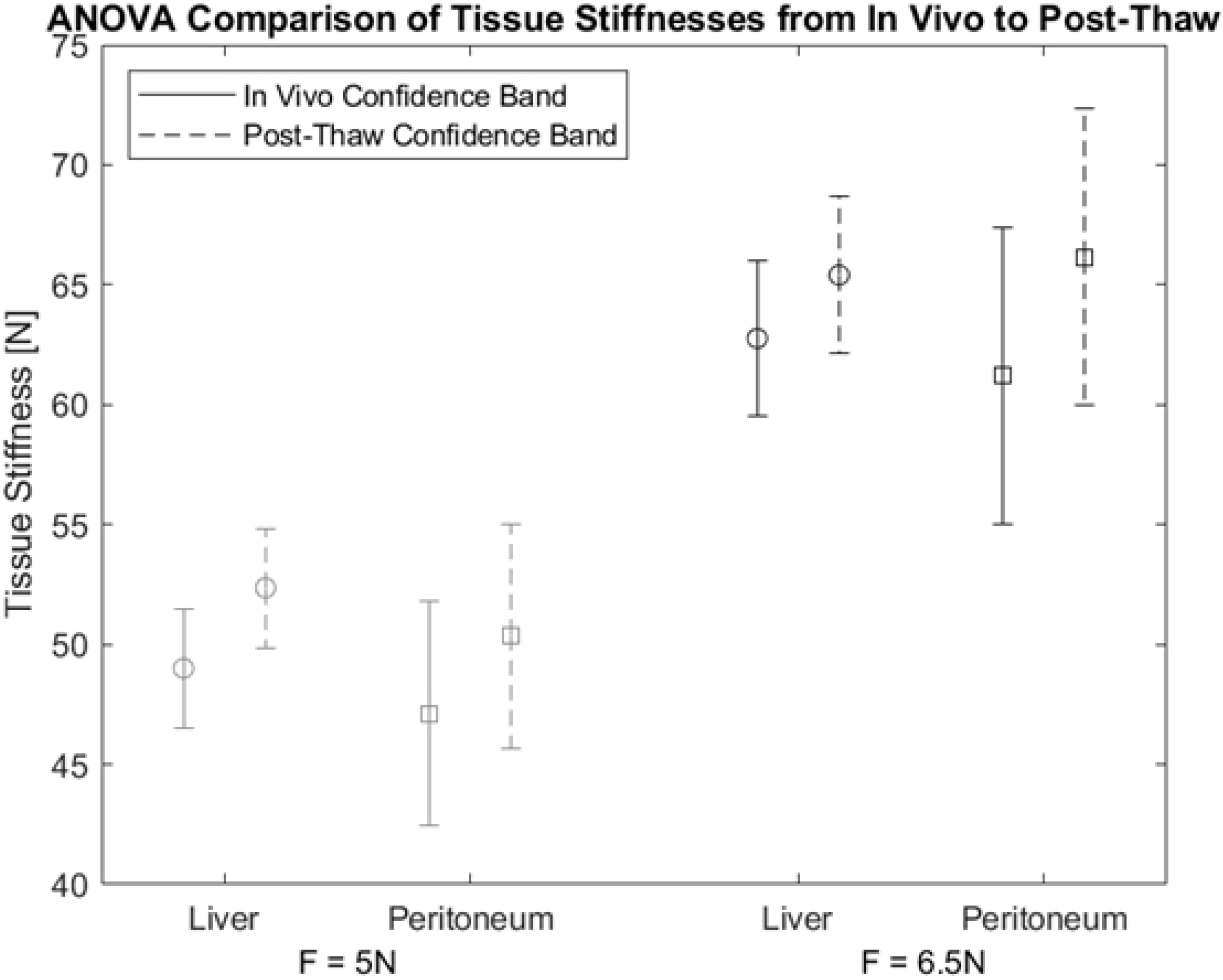
ANOVA comparison of Liver and Peritoneum from in vivo to post thaw, at both F=5N and F=6.5N. This plot shows the ANOVA confidence intervals (at p = 0.05) for the measured tissue stiffnesses from in vivo to post-thaw. Stiffnesses taken at lower force values are more faded (left), and higher force values are more saturated (right). The in vivo condition for each tissue is shown in solid lines, whereas the post-thaw condition is shown in dashed lines.

## Discussion

The measured change in stiffness of Liver tissues is statistically significant under the postmortem, ex vivo, and post-refrigeration states compared to the in vivo state; however, the change in stiffness under the post-thaw (preserved) state is not statistically significant. This indicates that the preservation process used in this study may affect liver and peritoneal stiffness, and cause it to revert back to in vivo levels. Thus, if the observations from this data were consistent for different tissues or possibly human tissues, existing data measured from preserved tissues may not greatly differ from similar characteristics of in vivo liver and peritoneum.

## Conclusions

This study findings provide evidence in support the use of preserved tissue to model realistic, in vivo tissue mechanical responses for medical training, at least for situations deemed adequately represented by porcine peritoneal and liver tissues. The study found no compelling evidence of large differences between in vivo and post thaw states to argue the contrary position: that in vivo data collection should be prioritized over post thaw approaches if the property of interest is stiffness. This is fortuitous, as preserved tissue tends to be easier to acquire and much easier to characterize for mechanical properties. These properties may be (and have been) used to create medical simulators, which are becoming increasingly common and relevant. Furthermore, the advent of “haptically-enhanced” virtual medical simulators (with force feedback) will would benefit from well-characterized tissue mechanical responses collected from such sources.

However, there are multiple limitations to this study. First, this study only considers a small sample population (n=5) of typically healthy, relatively young porcine models. Further, it targets the mechanical response of some tissues that are in no way representative of the complexity of animal physiology and its methodology is relatively agnostic to tissue thickness and surface conditions. There are other factors that affect the perception and handling experience of tissues, such as organ size, which may be altered after death. Other factors such as presence or volume of bodily fluids may also impact the experience of interacting with tissue.

Future work should analyze the effects of different preservation techniques on tissues, and consider more comprehensive testing on other tissues and tissue states – particularly tissues relevant to medical mannequins and medical simulators such as perfusion. Additionally, a robotically-actuated grasper could be used, using strain rates derived from this work to capture typical human grasp motions, in order to increase testing consistency while maintaining realistic human operating strain rates. This will also allow greater analysis of the intermediate tissue states, which did exhibit changes in stiffness that were statistically significant.

Furthermore, in vivo human data collection is necessary in order to characterize the variability in tissue properties within the same patient, and across different patients and evaluate whether the findings from porcine models generalize to humans, sustaining the conclusion that post-thaw data collection on suitably-preserved tissues may be justified in place of in vivo measurements. If this work found evidence of large differences between in vivo and post thaw states, it may have helped scientifically justify the added risk to conducting such research in humans. However, the lack of such evidence from this work does not help justify this increased risk.

## Data Availability

All data used as part of this research can be found in the Data Repository for University of Minnesota (DRUM) using the following link: https://conservancy.umn.edu/handle/11299/227135.

## Acknowledgements

The authors thank Drs. Paul Iazzo, Tinen Isles, and the staff at the Visible Heart Lab (VHL) at the University of Minnesota for providing valuable input, IACUC support, brokering animal access, and running the animal experiments in their facilities. The authors also acknowledge the input of Dr. Victor H. Barocas for discussion regarding analysis methodology (especially comparison at force levels or use of spherical indenters) and Mathew Kubala and Rebeca Smith for assisting in data collection.

## Supplementary Information

**S1 Fig. Measured Temperatures of Liver and Peritoneum Pre- and Post-Testing**.

